# Evaluation of Mental Health and Quality of Life among Afghan Refugees in Iran

**DOI:** 10.1101/2023.06.05.23290976

**Authors:** Mahdieh Saeidi, Saeed Sadr, Seyed Mehdi Samimi Ardestani, Sara Nooraeen

## Abstract

**Background:** Forced migration is significant in Middle Eastern countries. Afghanistan has a high rate of forced immigration due to decades of war and insecurity. This figure has risen, especially in the recent Taliban offensive. It is important to pay attention to the mental health of refugees and to evaluate the risk factors affecting the improvement of their mental health and quality of life. This study aims to investigate the mental health and quality of life among Afghan refugees living in Iran.

**Methods:** The study sample consisted of 200 Afghan refugees living in Tehran, Iran. Demographic characteristics, mental status and quality of life were investigated. One-sample t-test, chi-square, paired t-test, analysis of variance, and Scheffe post hoc test and logistic regression were used to analyze the data.

**Results:** Anxiety and insomnia were the most prevalent mental health symptoms among Afghan refugees. Depressive symptoms were higher in unemployed ones and women. Most of the participants (56.5%) had moderate quality of life and social relationship had the lowest score in quality-of-life domain.

**Conclusions:** The frequency of mental health disorders and low quality of life has been considerably high among Afghan Refugees in Iran. The mental health and quality of life of refugees should be further considered, especially after domestic wars and insecurities.

## Introduction

High quality of life and mental health status are two of the fundamental rights of every human, regardless of one’s race, religion, political inclination, economic situation, or social stance (King and Hinds 2011). This mobility should lead to a change of a person’s usual place of residence from origin or place of residence before migrating to a new destination or place of residence (Bonomi, Patrick et al. 2000). Immigration cannot always be considered a choice, and this situation in the case of emergency migration occurs due to factors that are beyond the will of the individual and are imposed on him/her (AG 2009). In recent years the number of forced Refugees has risen all around the world. According to United Nations High Commissioner for Refugees (UNHCR), more than 70 million people have currently been displaced (UNHCR 2019). Social crises such as violence, war, insecurity, poverty, and lack of social support are the main reasons for forced immigration (Steel, Chey et al. 2009, Alpak, Unal et al. 2015). The refugee crisis is more noticeable in Middle East countries. UNHCR reported that more than 500,000 people have already been made refugees by the Afghanistan crisis so far this year (UNHCR 2019). In 2019, the Taliban re-attack and the fall of the Afghanistan government, rising violence and insecurity have sparked a new wave of immigration to Iran and other neighboring countries. Therefore, the situation of Muslim Afghan refugees needs to be reconsidered.

Refugees face new stressors in the resettlement country (Nickerson, Bryant et al. 2011, Steel, Momartin et al. 2011). They experience different levels of homesickness, inequality, instability, financial problems, poor social support, and worry about the future in the resettled country (Steel, Silove et al. 2006, Steel, Chey et al. 2009, Miller and Rasmussen 2010). As well they might have the feeling of not belonging to the new place (Çelebi, Verkuyten et al. 2017). Hence, they could face various mental disorders in comparison with the general population. Several studies have evaluated the mental health condition of refugees in different nations (Fazel, Wheeler et al. 2005, Kessler, Petukhova et al. 2012, Slewa-Younan, Guajardo et al. 2015).

Edward Ng et al (Ng and Zhang 2020) reported that refugees are more likely to have mental health disorders in comparison to the Canadian-born population. A conducted systematic review by Marija Bogic et al (Bogic, Njoku et al. 2015) showed that mental health disorders are highly prevalent in war refugees even after many years. According to Shameran Slewa-Younan et al’s study (Slewa-Younan, Yaser et al. 2017), depression and post-traumatic stress disorder (PTSD) are high rates of disorders in Afghan refugees in Australia. In our country, Mohammadian et al (Mohammadian, Dadfar et al. 2005) showed that the prevalence of mental disorders among Afghan refugees in Tehran was 55.6%.

Considering the substantial increase in Afghan immigration to Iran, this study focuses on the quality of life and mental health condition of those who have migrated to Iran during the last four decades. Moreover, the second and third generation of Afghan refugees have also been considered.

## Materials and methods

This cross-sectional study was performed on 200 Afghan refugees older than 17 years old from March 2017 to March 2018 in Tehran, Iran. The method of data collection was to provide a questionnaire to Afghan citizens living in Tehran. In addition to providing a complete oral explanation of the purpose of the research, the conditions for collecting and maintaining the questionnaire, and the ethics of the research, during the review of the questionnaires, the participants in this study were reassured and asked to complete the questionnaire after careful study, if desired. All questionnaires in this study were conducted by a trained Afghan researcher and several interviewers. The use of the Afghan national interviewer greatly facilitated trust-building and better dialogue.

### 2.1. Study Design and Participants

Participants were selected by available sampling methods from Afghan population living in Tehran, Iran. Inclusion criteria were born in Afghanistan and for any reason at any age and in any circumstances, immigrated to Iran to continue living and have settled in the city of Tehran. We did not limit occupation, ethnicity, religion, length of stay, and reason for migrating. The population of Afghans born in Iran, those who could not speak Persian were excluded from the study. Afghan refugees living in other parts of the country were also excluded from the study, as well as those who did not wish to participate in completing the questionnaires. Also, due to the high level of illiteracy, for people who had a low level of education and were illiterate, each question was read to them by the questioner, to facilitate the completion of the questionnaire.

### 2.2. Instruments

1. A self-design demographic questionnaire was used to collect demographic characteristics such as gender, age, religion, ethnicity, level of education, income, marital status, living status (single-with family), and length of stay in Iran
2. The General Health Questionnaire – 28 (GHQ-28) is a self-report questionnaire to screen minor psychological disorders (Goldberg and Hillier 1979). This questionnaire was developed by Goldberg DP and Hillier VF. It has 4 sub-scales and each one has 7 questions. The reliability and validity of this questionnaire in Iran have been evaluated by S.M.R Taghavi (Taghavi 2008). In our study, we used this questionnaire to assess the mental health situation among the participants.
3. The Quality of Life Questionnaire of the World Health Organization (WHOQOL-BREF) (Abuse, Epidemiology et al. 2000) was developed by the WHOQOL Group. It is a cross-cultural quality of life assessment. This questionnaire has 4 domains which are: physical health (7 items), psychological health (6 items), social relationships (3 items), and environmental health (8 items). The validity and reliability of this instrument in Iran have been evaluated by S. Nedjat et al (Nedjat, Montazeri et al. 2008), this tool was used to evaluate the quality of life of the participants.

### 2.3. Statistical analysis

Quantitative variables were displayed by average and standard deviation and qualitative variables were displayed using frequency and percentage. One-sample t-test, chi-square, paired t-test, analysis of variance, and Scheffe post hoc test and logistic regression were used for evaluation of the mental health and quality of life and their relationship with demographic characteristics.

Statistical software SPSS version 21 was used for data analysis and the significance level for all tests was 0.005.

### 2.4. Ethical consideration

This study was performed after receiving the Ethical code from the ethical committee of Shahid Beheshti Medical University. Due to the legal issues regarding the departure of illegal refugees from the country and the fear arising from the legal issues by Afghan refugees, people were reassured that information would remain confidential, and surname registration or the exact address of the place of residence is not necessary.

Informed consent was obtained from all the participants. This research was approved by the ethics research committee of the Shahid beheshti University of Medial Sciences with unique number IR.SBMU.REC.1400.1147 and protocol number 22924. All the names were deleted to respect the anonymity of the participants.

## Results

### 3.1. Demographic Profile

Of 200 participants, 110 (55%) were male with a mean age of 34.37± 9.87 years (18-68). The average length of stay in Iran was 122.7± 108.8 months (from 3 months to 30 years). Table 1 shows the demographic characteristics in detail.

**Table 1.**
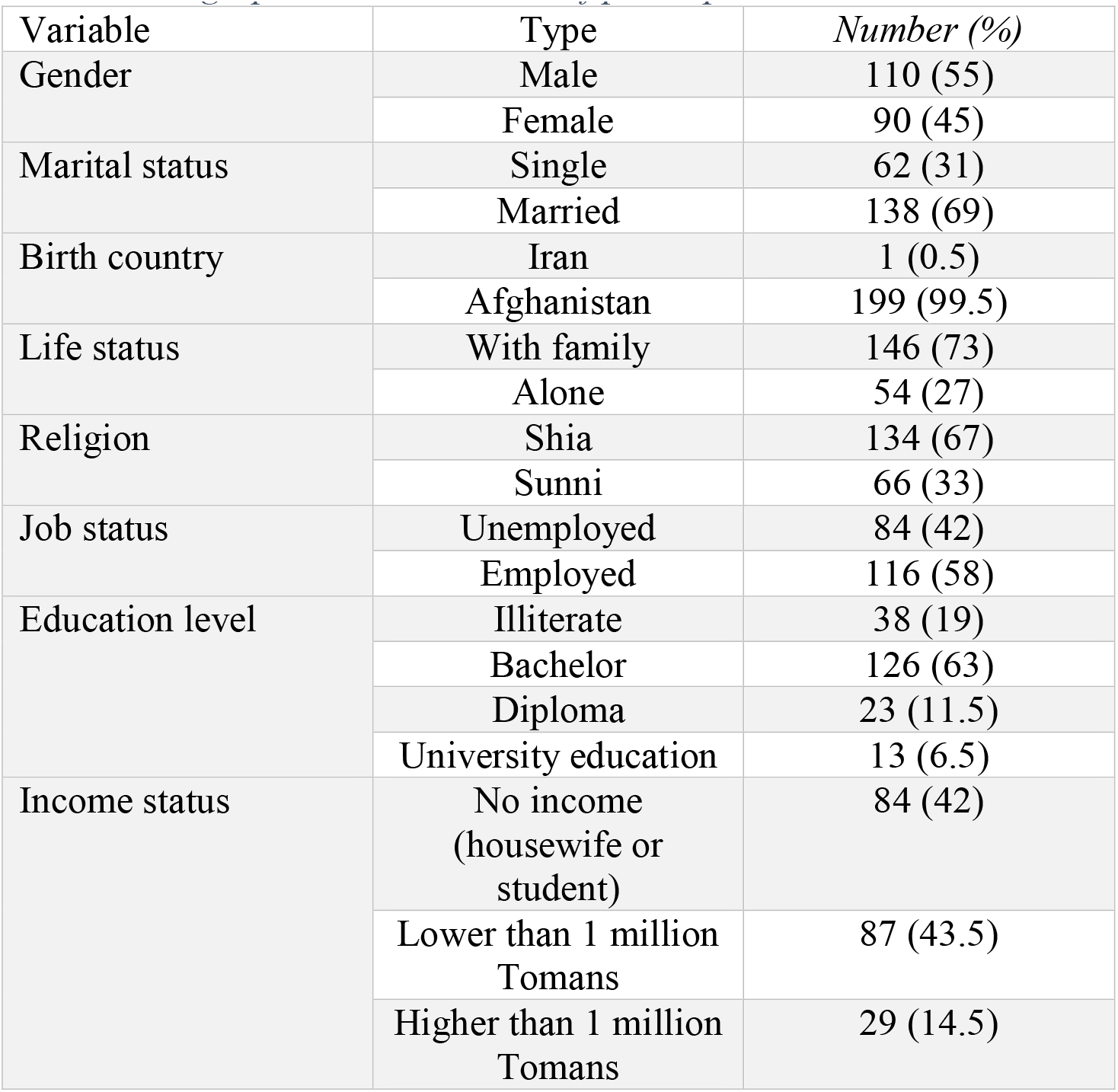
Demographic characteristics of participants

**Table 2.** Correlation matrix between fields of quality-of-life questionnaire

|  |  | Physical health | Psychological health | Social relationships | Environmental health |
| --- | --- | --- | --- | --- | --- |
| Physical health | P | 1 | 0.809 | 0.466 | 0.661 |
|  | P value |  | <0.001 | <0.001 | <0.001 |
| Psychological health | P | 0.809 | 1 | 0.501 | 0.640 |
|  | P value | <0.001 |  | <0.001 | <0.001 |
| Social | P | 0.466 | 0.501 |  | 0.587 |
| relationships | P value | <0.001 | <0.001 | 1 | <0.001 |
| Environmental health | P | 0.661 | 0.640 | 0.587 | 1 |
|  | P value | <0.001 | <0.001 | <0.001 |  |

*P: Pearson's coefficient of correlation*

### 3.2. Mental Health Status

We conducted the analysis based on the subscales of each questionnaire. The GHQ has 4 subscales: somatic symptoms (subscale A, items 1–7), anxiety and insomnia (B, 8–14), social dysfunction (C, 15–21), and severe depression (D, 22–28).

The mean score of the subjects in the severe depression (D) subscale was lower than other subscales and the mean score of the subjects in the anxiety and insomnia (B) was higher than other subscales. Comparing the mean score of each subscale to 6, which was the threshold, it was observed that only in subscale D, the mean score of individuals compared to the cut-off is significantly different (p = 0.097). The mean score in other subscales was significantly higher than the threshold of 6 (p <0.001). Also, in comparing the mean total score (38.64) of the threshold value of 22, it was observed that the mean total score was significantly higher than 22 (p <0.001).

Significant relationship was only seen between age increasing (p=0.006) and being married (p=0.012), with subscale A, age increasing and subscale B (p=0.053), length of stay in Iran and subscale C (p=0.003) and gender with subscale D (p<0.001). No significant relationship was observed between other demographic characteristics and mental health status in each GHQ subscale (p>0.05).

### 3.3. Quality of Life Status

Only 22.5% of participants had a good and very good quality of life. Others were moderate (56.5%), bad (19.5%) and very bad (1.5%). We analyzed the data based on 4 domains of the WHOQOL questionnaire: physical health, psychological health, social relationships, and environmental health. The mean score of social relationships was lower than other domains (40.38±22.04) and the mean score of physical health was higher than other domains (49.16±14.90). The mean score of other domains was 48.90±13.73 for psychological health and 48.20±12.47 for environmental health.

There was no statistically significant relationship between demographic characteristics and quality of life status in each WHOQOL domain (p>0.05) except for these:

The mean score of men in the psychological health domain was significantly higher than the mean score of women (p = 0.020), and the mean score of women in the social relationship’s domain was significantly higher than the mean score of men (p <0.001).

The mean score of Hazara people in the social relationship’s domain was significantly higher than the mean score of Tajik people (p = 0.002), also the mean score of Hazara people in the environmental health domain was higher than the average score of Tajiks. Statistically significant was marginally significant (p = 0.050).

The average score of single people in the physical health domain was significantly higher than the average score of married people (p = 0.001), also the average score of married people in the social relationship’s domain was significantly higher than the average score of single people (p<0.001).

The mean score of people living with family in social relationships was significantly higher than people living alone (p <0.001).

The average score of people who have a job was significantly higher than unemployed people in both physical (p=0.023) and psychological (0.001) dimensions.

There was a significant positive correlation between length of stay and social relationships (r = 0.299, p <0.001) so that with the increasing length of stay, social relations were more.

## Discussion

This study aimed to evaluate the mental health status and quality of life among Afghan refugees in Iran. As regards the increasing rate of Afghan immigration to neighboring countries, especially Iran, detecting their mental health problems is of value.

The results of our study showed that the mean score of GHQ is above the normal (mean score=38.64, p= 0.001) and anxiety and insomnia (subscale B) are the most prevalent health conditions among Afghan refugees. There are a lot of studies that addressed the mental health status of refugees. Farah Ahmad et al showed that PTSD is one of the most common mental health disorders among Afghan refugees in Canada and it is significantly correlated with age, unemployment, social support, and self-rated health, and higher social support was significantly associated with a lower rate of PTSD (Ahmad, Othman et al. 2020). A systematic review by Qais Alemi et al demonstrated that depression and PTSD have moderate to severe prevalence among Afghan refugees (Alemi, James et al. 2014).

In our study, anxiety and insomnia (subscale B) are the most common symptoms in the participants. Higher age is significantly associated with the higher symptoms of anxiety and insomnia. The chance of anxiety symptoms increased by 7% with each year of age (OR=1.066, 95%CI (0.99-1.14)), and this association is marginally significant (p=0.503).

Refugees are faced with numerous stressors in the resettled country (Matheson, Jorden et al. 2008). One of the influential factors in this issue could be the experience of discrimination in the resettled country. According to Qais Alemi and Carl Stempel, discrimination is significantly associated with distress, and previous traumatic experiences exacerbate this association (Alemi and Stempel 2018). Sleep disorder is also common in refugees (Richter, Baumgärtner et al. 2020, Lies, Jobson et al. 2021). MPsych (Clin) et al reported that of 2703 refugees in their study, 75.5% had moderate to severe sleep disorders (Lies, Mellor et al. 2019). Pre-migration traumatic experiences could be another factor in mental health conditions. In LeMaster, J W et al’s study, depressive and PTSD symptoms are higher in refugees who experienced more traumatic events before immigration (LeMaster, Broadbridge et al. 2018). On the other hand, help-seeking for mental health conditions is less among refugees (Hinton, Howes et al. 2008, Andrade, Alonso et al. 2014, Clement, Schauman et al. 2015). This could be for the following reasons: cultural issues and stigma, structural barriers such as financial problems and instability, and specific barriers such as mistrust and lack of confidentiality (Byrow, Pajak et al. 2020).

Somatic symptoms are common in refugees (Çelebi, Verkuyten et al. 2017, Khan and Amatya 2017, Oda, Tuck et al. 2017), our study showed that in the field of somatic symptoms (subscale A), high age (p=0.006) and being married (p=0.012) are the two factors associated with the score. It could be due to the association between aging and disease (Biran, Zada et al. 2017, Pagiatakis, Musolino et al. 2021). A. M. Gerritsen et al in a conducted study on 178 refugees and 232 asylum seekers showed that physical and mental health disorders are highly common in the refugees and about half of the participants suffered from at least one chronic disorder, depression, and anxiety, and higher age was correlated with a poor general health condition and chronic disorders (Gerritsen, Bramsen et al. 2006). Married people, on the other hand, are generally older than single people, which may explain the association between marital status and somatic symptoms.

In the field of social dysfunction (subscale C) increased length of stay was significantly associated with a decrease in social dysfunction (p=0.003). Our findings showed that one-month increase in length of stay is associated with a 1% reduction in social dysfunction (OR=0.995, 95%CI (0.991-0.998); p=0.003)). Although refugee problems can persist for years after migration, adapting to the new environment and finding a job can help reduce stress and improve the mental health status (Berry 1997). The longer the resettlement process in the new country takes, the greater the likelihood of future mental disorders (Laban, Gernaat et al. 2004). It seems that increasing the number of years of stay increases adaptation to the new environment. On the other hand, social support correlates with a lower rate of mental disorders (Schwarz-Nielsen and Elklitt 2009, Elklit, Lasgaard et al. 2012).

In the severe depression field (subscale A), there was a significant association between female gender (68.4% female vs. 31.6% male) and unemployment (55.1% unemployment vs. 44.9% employment) with a higher rate of depressive symptoms (p< 0.001). In many studies, female gender and unemployment have been suggested as predictors of depression. In a study by Sharp MB et al the prevalence of depression was 55.9% among 272 female refugees and having an unemployed spouse was a risk factor for higher levels of depression (Sharp, Parpia et al. 2020). In another study by A. Ceren on 781 Syrian refugees, the prevalence of depression was 37.4%, and being female was a predictor for depression (OR=5.1). C Hawkes et al reported that high levels of depressive symptoms were seen in 21.2% of 66 afghan refugees residing in Australia and were more prevalent among female participants (41.2%, 95% CI: 18.4%-67.1%) (Hawkes, Norris et al. 2021). Unemployment seems to be causing depressive symptoms by increasing financial problems. Having an unemployed spouse and worries about children and future life could be some causes of higher rate of depressive symptoms among women.

Our study showed that only 22.5% of participants had a good and very good quality of life and the lowest score of quality-of-life domains was for social relationships (40.38± 22.04). One of the main problems of refugees in the new country is the lack of adequate social support, which can affect their social relations and quality of life. On the other hand, the experience of discrimination and the feeling of not belonging to the resettled country could affect their social relationships. In our study, women, married, living with family, and more years of living in Iran were the factors attributed to better social relationships. It seems that familial support and adaptation to the new country could help refugees to have better social relations (Lamba and Krahn 2003, Busch Nsonwu, Busch-Armendariz et al. 2013, McGregor, Melvin et al. 2015). Like the GHQ results, women have lower levels of psychological health in comparison to men (46.86 vs. 51.39, p=0.020). On the other hand, male refugees who have a job have a higher quality of life than the unemployed ones. These findings show the importance of paying more attention to women (Awaad, Fisher et al. 2019) and creating job opportunities to increase the quality of life of refugees.

Our study shed some light on the mental health and quality of life of Afghan refugees in Iran. Our results showed that Afghan refugees in Iran need special attention in terms of mental health and quality of life. Providing facilities such as access to mental health services, job creation, and support for the unemployed and women, coping skills training and public awareness of non-discrimination should be on the agenda. In past years, with the re-attack of the Taliban and the fall of the Afghanistan government, there has been a new wave of displacement and migration of Afghans to neighboring countries, including Iran. Therefore, the mental state and quality of life of Afghan refugees need to be re-examined. More studies on a large population of refugees are suggested.

It is necessary to think more about improving the situation and to try to create a platform for the prevention and treatment of existing cases and to remove the obstacles by programming. Because, by solving these problems, it is possible to raise refugees who are more satisfied with their quality of life and have good mental health, with the hope of raising people who are more capable of performing their duties as part of the family and society.

### Limitations

Small number of participants, legal issues regarding the departure of illegal refugees and also considering that most of the population studied were illiterate or low-educated, therefore, the information received is likely to be distorted and the results obtained could not be generalized to the entire population of Afghan refugees.

## Data Availability

All data produced in the present study are available upon reasonable request to the authors

## References

Abuse, N. I. o. D., et al. (2000). National household survey on drug abuse: Main findings, National Institute on Drug Abuse, Division of Epidemiology and Statistical ….

AG, A. (2009). “[Imigrants movements in iran from 1994-2004] “ jamiat.

Ahmad, F., et al. (2020). “Posttraumatic stress disorder, social support and coping among Afghan refugees in Canada.” Community mental health journal 56(4): 597–605.

Alemi, Q., et al. (2014). “Psychological distress in Afghan refugees: A mixed-method systematic review.” Journal of immigrant and minority health 16(6): 1247–1261.

Alemi, Q. and C. Stempel (2018). “Discrimination and distress among Afghan refugees in northern California: The moderating role of pre-and post-migration factors.” PloS one 13(5): e0196822.

Alpak, G., et al. (2015). “Post-traumatic stress disorder among Syrian refugees in Turkey: a cross-sectional study.” International journal of psychiatry in clinical practice 19(1): 45–50.

Andrade, L. H., et al. (2014). “Barriers to mental health treatment: results from the WHO World Mental Health surveys.” Psychological medicine 44(6): 1303–1317.

Awaad, R., et al. (2019). “Development and validation of the Muslims’ perceptions and attitudes to mental health (M-PAMH) scale with a sample of American Muslim women.” Journal of Muslim Mental Health 13(2).

Berry, J. W. (1997). “Immigration, acculturation, and adaptation.” Applied psychology 46(1): 5–34.

Biran, A., et al. (2017). “Quantitative identification of senescent cells in aging and disease.” Aging cell 16(4): 661–671.

Bogic, M., et al. (2015). “Long-term mental health of war-refugees: a systematic literature review.” BMC international health and human rights 15(1): 1–41.

Bonomi, A. E., et al. (2000). “Validation of the United States’ version of the world health organization quality of life (WHOQOL) instrument.” Journal of clinical epidemiology 53(1): 1–12.

Busch Nsonwu, M., et al. (2013). “Marital and familial strengths and needs: refugees speak out.” Journal of Ethnic And Cultural Diversity in Social Work 22(2): 129–144.

Byrow, Y., et al. (2020). “Perceptions of mental health and perceived barriers to mental health help-seeking amongst refugees: A systematic review.” Clinical psychology review 75: 101812.

Çelebi, E., et al. (2017). “Ethnic identification, discrimination, and mental and physical health among Syrian refugees: The moderating role of identity needs.” European journal of social psychology 47(7): 832–843.

Clement, S., et al. (2015). “What is the impact of mental health-related stigma on help-seeking? A systematic review of quantitative and qualitative studies.” Psychological medicine 45(1): 11–27.

Elklit, A., et al. (2012). “Social support, coping and posttraumatic stress symptoms in young refugees.” Torture: quarterly journal on rehabilitation of torture victims and prevention of torture 22(1): 11–23.

Fazel, M., et al. (2005). “Prevalence of serious mental disorder in 7000 refugees resettled in western countries: a systematic review.” The Lancet 365(9467): 1309–1314.

Gerritsen, A. A., et al. (2006). “Physical and mental health of Afghan, Iranian and Somali asylum seekers and refugees living in the Netherlands.” Social psychiatry and psychiatric epidemiology 41(1): 18–26.

Goldberg, D. P. and V. F. Hillier (1979). “A scaled version of the General Health Questionnaire.” Psychological medicine 9(1): 139–145.

Hawkes, C., et al. (2021). “Resettlement stressors for women of refugee background resettled in regional Australia.” International journal of environmental research and public health 18(8): 3942.

Hinton, D. E., et al. (2008). “Toward a medical anthropology of sensations: Definitions and research agenda.” Transcultural Psychiatry 45(2): 142–162.

Kessler, R. C., et al. (2012). “Twelve□month and lifetime prevalence and lifetime morbid risk of anxiety and mood disorders in the United States.” International journal of methods in psychiatric research 21(3): 169–184.

Khan, F. and B. Amatya (2017). “Refugee health and rehabilitation: challenges and response.” Journal of rehabilitation medicine 49(5): 378–384.

King, C. R. and P. S. Hinds (2011). Quality of life: from nursing and patient perspectives, Jones & Bartlett Publishers.

Laban, C. J., et al. (2004). “Impact of a long asylum procedure on the prevalence of psychiatric disorders in Iraqi asylum seekers in The Netherlands.” The Journal of nervous and mental disease 192(12): 843–851.

Lamba, N. K. and H. Krahn (2003). “Social capital and refugee resettlement: The social networks of refugees in Canada.” Journal of International Migration and Integration/Revue de l’integration et de la migration internationale 4(3): 335–360.

LeMaster, J. W., et al. (2018). “Acculturation and post-migration psychological symptoms among Iraqi refugees: A path analysis.” American Journal of Orthopsychiatry 88(1): 38.

Lies, J., et al. (2021). “Postmigration stress and sleep disturbances mediate the relationship between trauma exposure and posttraumatic stress symptoms among Syrian and Iraqi refugees.” Journal of Clinical Sleep Medicine 17(3): 479–489.

Lies, J., et al. (2019). “Prevalence of sleep disturbance and its relationships with mental health and psychosocial issues in refugees and asylum seekers attending psychological services in Australia.” Sleep health 5(4): 335–343.

Matheson, K., et al. (2008). “Relations between trauma experiences and psychological, physical and neuroendocrine functioning among Somali refugees: Mediating role of coping with acculturation stressors.” Journal of immigrant and minority health 10(4): 291–304.

McGregor, L. S., et al. (2015). “Familial separations, coping styles, and PTSD symptomatology in resettled refugee youth.” The Journal of nervous and mental disease 203(6): 431–438.

Miller, K. E. and A. Rasmussen (2010). “War exposure, daily stressors, and mental health in conflict and post-conflict settings: bridging the divide between trauma-focused and psychosocial frameworks.” Social science & medicine 70(1): 7–16.

Mohammadian, M., et al. (2005). “Screening for mental disorders among Afghan immigrants residing in Tehran.” Iranian Journal of Psychiatry and Clinical Psychology 11(3): 270–277.

Nedjat, S., et al. (2008). “Psychometric properties of the Iranian interview-administered version of the World Health Organization’s Quality of Life Questionnaire (WHOQOL-BREF): a population-based study.” BMC health services research 8(1): 1–7.

Ng, E. and H. Zhang (2020). “The mental health of immigrants and refugees: Canadian evidence from a nationally linked database.” Health Rep: 3–12.

Nickerson, A., et al. (2011). “The familial influence of loss and trauma on refugee mental health: A multilevel path analysis.” Journal of traumatic stress 24(1): 25–33.

Oda, A., et al. (2017). “Health care needs and use of health care services among newly arrived Syrian refugees: a cross-sectional study.” CMAJ open 5(2): E354.

Pagiatakis, C., et al. (2021). “Epigenetics of aging and disease: a brief overview.” Aging clinical and experimental research 33(4): 737–745.

Richter, K., et al. (2020). “Sleep disorders in migrants and refugees: a systematic review with implications for personalized medical approach.” The EPMA journal 11(2): 251.

Schwarz-Nielsen, K. H. and A. Elklitt (2009). “An evaluation of the mental status of rejected asylum seekers in two Danish asylum centers.” Torture 19(1): 51–59.

Sharp, M., et al. (2020). “Prevalence of and risk factors for depression among female Syrian refugees and Jordanians with chronic disease: a pilot study.” East Mediterr Health J 26.

Slewa-Younan, S., et al. (2015). “A systematic review of post-traumatic stress disorder and depression amongst Iraqi refugees located in western countries.” Journal of immigrant and minority health 17(4): 1231–1239.

Slewa-Younan, S., et al. (2017). “The mental health and help-seeking behaviour of resettled Afghan refugees in Australia.” International journal of mental health systems 11(1): 1–8.

Steel, Z., et al. (2009). “Association of torture and other potentially traumatic events with mental health outcomes among populations exposed to mass conflict and displacement: a systematic review and meta-analysis.” Jama 302(5): 537–549.

Steel, Z., et al. (2011). “Two year psychosocial and mental health outcomes for refugees subjected to restrictive or supportive immigration policies.” Social science & medicine 72(7): 1149–1156.

Steel, Z., et al. (2006). “Impact of immigration detention and temporary protection on the mental health of refugees.” The british journal of psychiatry 188(1): 58–64.

Taghavi, M. R. (2008). “The normalization of general health questionnaire for Shiraz University students (GHQ-28).” Daneshvar raftar 15(28): 1–12.

UNHCR (2019). “UNHCR.” The UN Refugee Agency, Figures at a glance.

